# Investigating the causal relationship between allergic disease and mental health

**DOI:** 10.1101/2021.04.28.21256258

**Authors:** Ashley Budu-Aggrey, Sally Joyce, Neil M Davies, Lavinia Paternoster, Marcus R. Munafò, Sara J Brown, Jonathan Evans, Hannah M. Sallis

**Affiliations:** Medical Research Council (MRC) Integrative Epidemiology Unit at the University of Bristol, Bristol, UK; Bristol Medical School, Population Health Sciences, University of Bristol, Bristol, UK; K.G. Jebsen Center for Genetic Epidemiology, Department of Public Health and Nursing, NTNU, Norwegian University of Science and Technology, Norway; School of Psychological Science, University of Bristol, Bristol, UK; Centre for Genomic and Experimental Medicine, Institute of Genetics and Molecular Medicine, The University of Edinburgh, Edinburgh, UK; Centre for Academic Mental Health, Population Health Sciences, University of Bristol, Bristol, UK

**Keywords:** allergic disease, association, asthma, causal, atopic dermatitis, hay fever, Mendelian randomization, mental health

## Abstract

**Background:** Observational studies have reported an association between allergic disease and mental health, but a causal relationship has not been established.

**Objective:** To use Mendelian Randomization (MR) to investigate a possible causal relationship between atopic disease and mental health phenotypes.

**Methods:** The observational relationship between allergic disease and mental health was investigated in UK Biobank. The direction of causality was investigated with bidirectional two-sample MR using summary-level data from published genome-wide association studies. A genetic instrument was derived from associated variants for a broad allergic disease phenotype to test for causal relationships with various mental health outcomes. Genetic instruments were also derived for mental health conditions to assess causality in the reverse direction. We also investigated if these relationships were specific to atopic dermatitis (AD), asthma or hay fever.

**Results:** The broad allergic disease phenotype was phenotypically associated with most measures of mental health, but we found little evidence of causality in either direction. However, we did find evidence of genetic liability for bipolar disorder causally influencing hay fever risk (OR=0.94 per doubling odds of bipolar disorder risk; 95%CI=0.90-0.99; *P*-value=0.02), but evidence of a phenotypic association was weak.

**Conclusions:** Few of the phenotypic associations between allergic disease and mental health were replicated. Any causal effects we detected were considerably attenuated compared to the phenotypic association. This suggests that most co-morbidity observed clinically is unlikely to be causal.

**Clinical Implication:** We found little evidence that genetic predisposition to allergic disease *causes* mental ill-health, and *vice versa*, which suggests that intervening to prevent onset of allergic disease is unlikely to directly prevent the onset of mental ill-health.

**Key Messages:**

- Mendelian randomization effect estimates suggest that the phenotypic association between allergic disease and mental health is likely to be inflated
- Causal analysis was unable to corroborate the phenotypic associations observed between allergic disease and mental health phenotypes
- Intervening on an individual’s allergic disease is not likely to directly improve their mental health

**Capsule summary:** Mendelian Randomization suggests that evidence of a causal relationship between allergic disease and mental health phenotypes is weak. It is unlikely that intervening to prevent onset of allergic disease will prevent poor mental health.

## Introduction

There is a well-documented relationship between allergic disease (including asthma, atopic dermatitis (AD) and hay fever) and mental health. A recent population-based study in Taiwan reported an up to 2-fold risk of allergic disease sufferers developing psychiatric disorders^1^. However, it is unclear whether this association is causal, or whether confounding factors, or reverse causality, could explain the observed association. Prevalence of common mental health disorders and allergic disease is increasing^2^. Common mental health disorders such as anxiety and depression are some of the largest contributors to the global burden of disease. Depression is currently the leading cause of disability worldwide with around 4.4% of the world population suffering^3^, and depressive disorders accounting for 44 million lost years of productive life^4^. Establishing whether there is a causal relationship is therefore important, as this could highlight whether effectively treating allergic disease would lead to a reduction in the burden of mental health issues (or vice versa).

Increased prevalence of disorders such as depression, anxiety, schizophrenia, conduct disorder, and autism are observed among individuals with AD, particularly those most severely affected^5,6^. Other studies have found evidence of an association between asthma and hay fever with bipolar disorder, depression and schizophrenia^7,8^. Several hypotheses have been suggested for a causal role of allergic disease on mental health; these include both psychosocial and biological mechanisms. It is possible that social consequences of allergic disease, such as embarrassment due to visible lesions or itching resulting from AD, or results of sleep deprivation are the driving force behind later mental health issues. An alternative hypothesis is the ‘inflammatory hypothesis’, which suggests that the effects of allergic disease on the immune system (such as disturbances in the inflammatory system or increased levels of inflammatory cytokines) could contribute to the presence of mental health disorders^9,10^.

If these associations reflect a true causal effect of allergic disease on mental health, this could suggest targets for potential intervention and prevention targets for subsequent mental health problems. For example, screening, monitoring and/or early intervention among allergic disease patients could reduce the risk of later bipolar disorder. If the effects do act via inflammatory mechanisms, then repurposing existing treatments for inflammatory disease could be effective here. Nonsteroidal anti-inflammatory drugs (NSAIDS) have been suggested to be an effective adjunct therapy for bipolar disorder, however this finding remains inconclusive^11,12^. Studies investigating the progression of disease rather than onset will further inform on the effectiveness of repurposing opportunities to treat current sufferers^13^.

We used data from the UK Biobank study to further investigate the phenotypic associations between allergic disease and mental health in a population of European ancestry. We also performed two-sample Mendelian randomization (MR) using summary data from relevant genome-wide association studies (GWAS) to investigate the causal nature of these associations. MR is an approach that enables us to infer causality using observational data. The method uses genetic variants as a proxy for modifiable exposures, and subject to the instrumental variable assumptions holding, should not be subject to the issues affecting conventional observational epidemiology^14,15^. This is based on the assumptions that the instrumental variable is truly associated with the exposure and not associated with confounders, and also has an effect upon the outcome via the exposure and not via alternative pathways^14^.

Disentangling the nature of the relationship between allergic disease and mental health could enable us to focus efforts on improving intervention or prevention strategies. Whether there is a causal effect of allergic disease on mental health, as suggested by previous observational studies, is a critical question for intervening on the initial presentation of allergic disease to improve mental health outcomes of patients.

## Methods

### Study populations

#### Phenotypic association – UK Biobank

Data was available from the UK Biobank for individuals aged between 37 and 73 years^16^, including those with asthma, AD, hay fever and the broad allergic disease phenotype (any one of asthma, AD or hay fever) (Table 1). Phenotype data was also available for a number of mental health and personality traits including depression, anxiety, bipolar disorder, schizophrenia, stress and neuroticism. All individuals were of European ethnicity who had provided written informed consent. UK Biobank is approved by the National Health Service National Research Ethics Service (ref 11/NW/0382; UK Biobank application number 9142).

**Table 1.**
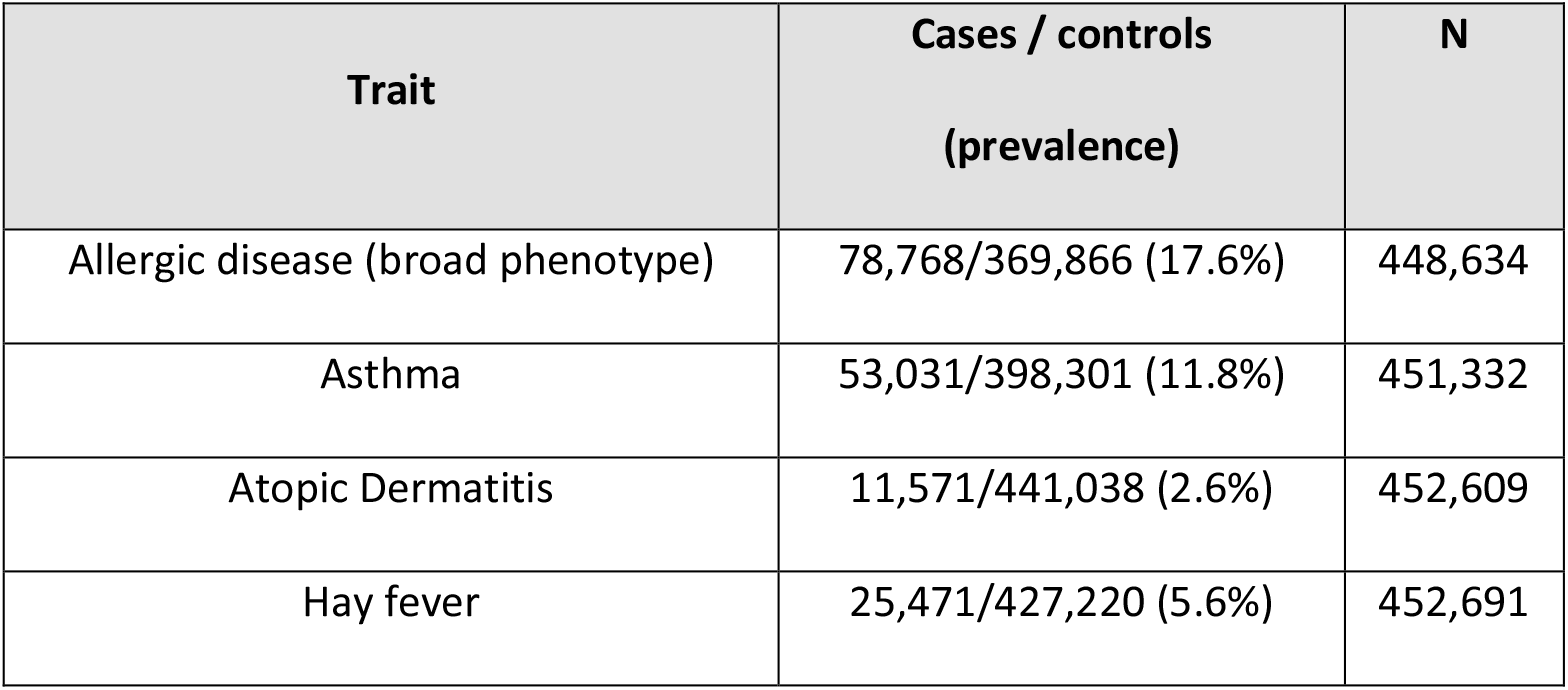
Descriptive statistics of UK Biobank individuals with allergic disease.

| Trait | Cases / controls<br>(prevalence) | N |
| --- | --- | --- |
| Allergic disease (broad phenotype) | 78,768/369,866 (17.6%) | 448,634 |
| Asthma | 53,031/398,301 (11.8%) | 451,332 |
| Atopic Dermatitis | 11,571/441,038 (2.6%) | 452,609 |
| Hay fever | 25,471/427,220 (5.6%) | 452,691 |

##### Mental health and personality phenotypes

Self-reported measures of depression, anxiety, bipolar disorder, schizophrenia and stress were derived from the non-cancer illness item asked at the first assessment centre (field ID: 20002). Diagnoses of major depressive disorder (MDD) and bipolar disorder were derived from responses to the mental health touchscreen questionnaire completed at the first assessment centre. Neuroticism summary scores were based on 12 neurotic behaviour domains as derived by Smith et al^17^.

##### Allergic disease phenotypes

Asthma, AD and hay fever phenotypes were also derived from the self-reported non-cancer illness item at the first assessment centre. Participants were designated as controls for the relevant phenotype if they did not report asthma, AD or hay fever based on the self-reported non-cancer illness item, and if they did not report a doctor diagnosis of asthma or AD/hay fever (field ID: 6152). A broad allergic disease phenotype was derived from these phenotypes, participants reporting either asthma, hay fever or AD (or any combination of these phenotypes) were assigned case status. Participants who did not report either asthma, AD or hay fever as described for the individual phenotypes were designated as controls.

#### Causal relationship - Summary GWAS data

Published summary GWAS results were available for the most recent GWAS for the broad allergic disease phenotype (n=360,868), which had considered the existence of any one of the atopic triad (asthma, AD and hay fever)^18^. Summary GWAS results were also available for asthma (n=127,669)^19^, hay fever (n=38,838)^20^, and AD (n=40,835)^21^. Similarly, published GWAS results were available for the mental health outcomes investigated; major depressive disorder (MDD)^22^, bipolar disorder^23^, schizophrenia^24^, neuroticism^25^ and anxiety^26^. All published results were from analyses restricted to individuals of European ancestry.

### Statistical analyses

#### Observational analyses

Observational analyses were carried out using individual level data from UK Biobank participants to investigate the phenotypic association between the allergic disease phenotypes and psychiatric traits. This was performed using linear or logistic regression as appropriate while adjusting for age and sex. All observational analyses were performed using Stata 15^27^. Given the multi-testing burden, we note that a global *P*=0.05 is equivalent to an individual test *P*-value of 0.0016 (32 tests).

#### Analysis of causal relationships

A genetic instrument was defined for the broad allergic disease phenotype using 89 independent (r^2^ < 0.01) single-nucleotide polymorphisms (SNPs) from loci reported to be most strongly associated (*P* < 5×10^−8^) by Ferreira and colleagues^18^. Genetic instruments were also defined for asthma (16 SNPs) ^19^, AD (23 SNPs) ^21^ and hay fever (37 SNPs)^20^ using independent (r^2^ < 0.01) SNPs associated at the genome-wide significance threshold (*P*<5×10^−8^) from the largest GWAS of these conditions (see Tables E1 to E4 in the Online Repository). Effect sizes for each of these SNPs were extracted from recent GWAS of each mental health phenotype to create our outcome datasets. MR was then performed using the TwoSample MR R package^28^ to determine the causal effect of allergic disease liability upon the risk for mental disorders (Figure 1a). A causal estimate was obtained using the inverse-variance weighted (IVW) method, which is akin to a weighted regression of the SNP-outcome coefficients upon the SNP-exposure coefficients. In doing so the SNP-exposure and SNP-outcome associations were combined in a meta-analysis. For ease of interpretation, the resulting causal estimates were multiplied by 0.693 to represent the change in outcome per doubling odds of the exposure as demonstrated by Burgess and Labreque^29^.

**Figure 1.**
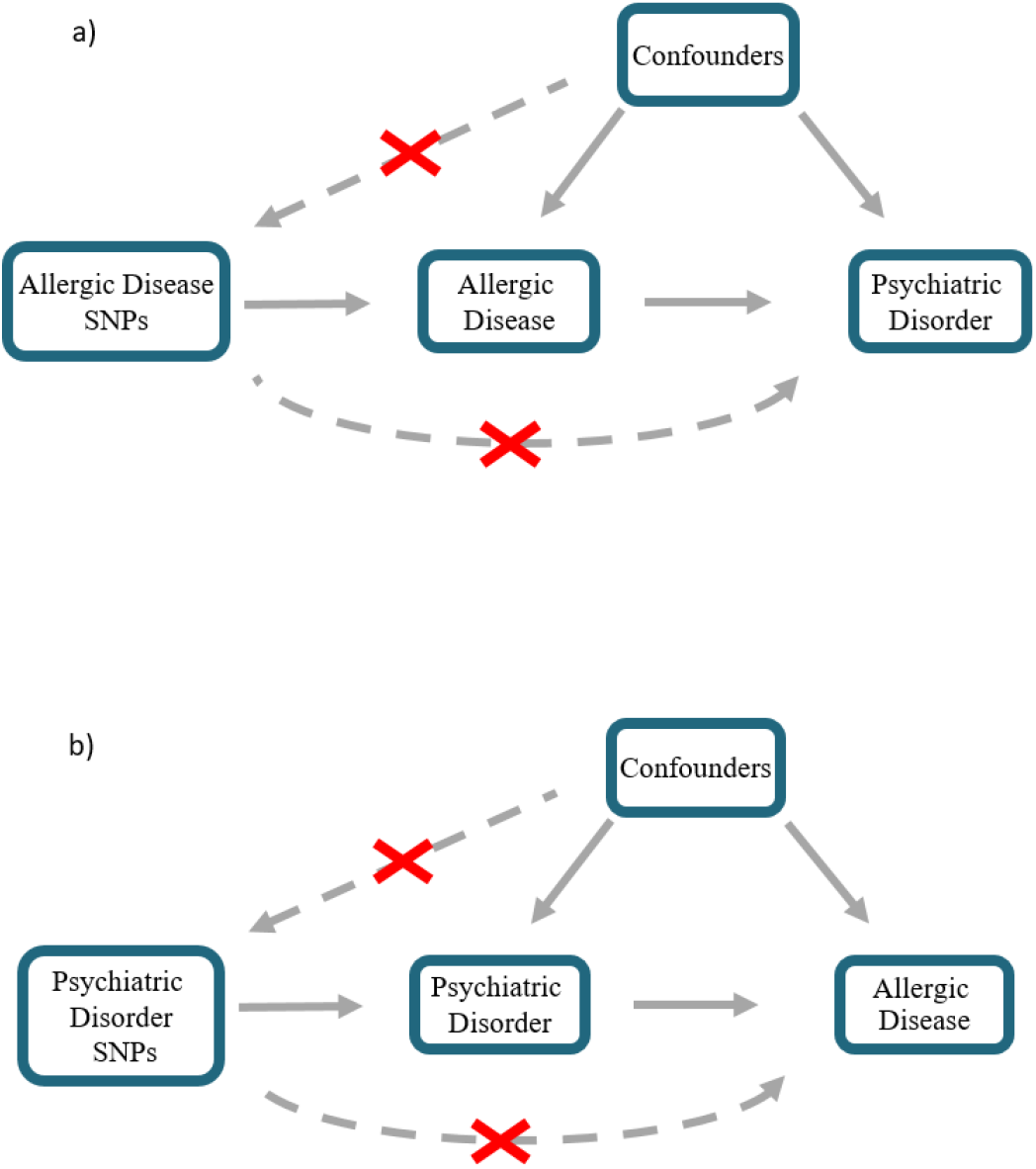
Schematic representation of MR analyses. a) Allergic disease SNPs were used as genetic instruments to investigate the causal effect of allergic disease liability upon various psychiatric disorders and mental health outcomes. b) SNPs for various psychiatric disorders were used as genetic instrument to investigate to causal effect of liability for psychiatric disorders and mental health outcomes upon allergic disease. SNP = single nucleotide polymorphism

MR analyses were also performed in the reverse direction to estimate the causal effect of liability for mental health traits upon allergic disease risk (Figure 1b). Genetic instruments were derived from associated independent variants (*P* < 5×10^−8^; r^2^ < 0.01) reported for MDD (32 SNPs)^22^, bipolar disorder (22 SNPs)^23^, anxiety (5 SNPs)^30^, schizophrenia (79 SNPs)^24^ and neuroticism (66 SNPs)^25^ (see Tables E5 to E9 in the Online Repository).

#### Sample overlap

UK Biobank participants were included in the discovery samples for the broad allergic disease phenotype, hay fever, MDD and anxiety. This sample overlap has the potential to bias causal effect estimates towards the phenotypic exposure-outcome association^31^. To avoid sample overlap, we generated genetic instruments for MDD (Wray et al. 2018) based on summary statistics excluding UK Biobank (31 SNPs). Summary data for hay fever excluding UK Biobank was also available when hay fever was the outcome^20^. An alternative anxiety genetic instrument was also generated using SNP-exposure coefficients from an older anxiety GWAS that did not include UK Biobank^26^. We do note that due to limitations of data availability, there were some analyses where sample overlap could not be avoided, these being when investigating causality between the broad allergic disease and neuroticism (estimated sample overlap = 42.0%) as well as for hay fever and neuroticism (estimated sample overlap = 44.4%).

#### Sensitivity analyses

The causal estimate could be biased by SNPs within the genetic instrument which act through a horizontally pleiotropic pathway (whereby a SNP affects the outcome by a path other than via the exposure – the bottom dotted arrows in Fig 1). Various sensitivity analyses have been designed to detect such pleiotropy and ensure that the assumption that instruments are only influencing the outcome via the exposure is not violated. We used four sensitivity methods that all rely on different assumptions to test for or account for potential pleiotropy. These are MR-Egger regression^32^, weighted median analysis^32^ and the weighted mode-based estimate (MBE)^33^ and Cochran’s Q statistic^34^. In addition, the Steiger directionality test was also performed to ensure that the variance explained by the genetic instrument was greater in the exposure compared to the outcome^35^.

All MR analyses were performed using R (www.r-project.org). The code and datasets used to carry out the MR analyses are available on GitHub (https://github.com/abudu-aggrey/Allergic_Disease_Mental_Health_MR).

## Results

### Observational analyses

We found strong observational evidence for an association of the broad allergic disease phenotype with both self-reported (OR = 1.45; 95% CI = 1.41, 1.50; *P*-value = 3.63×10^−130^) and clinically diagnosed depression (OR = 1.40; 95% CI = 1.35, 1.46; *P*-value = 2.63×10^−76^) (Figure 2a). The direction and magnitude of the effect estimate is consistent with that previously reported in a Taiwanese population ^1^ suggesting an increased odds of depression in allergic disease sufferers compared to non-sufferers. A strong association was also seen between depression and the individual asthma, AD and hay fever phenotypes, though with different magnitudes (Figure 2a, see Table E11 in the Online Repository). However, we do note that the adult prevalence of AD (5-10%)^36^, asthma (18.2%)^37^ and hay fever (10-30%)^38^ in the population is greater than that identified in UK Biobank (Table 1).

**Figure 2.**
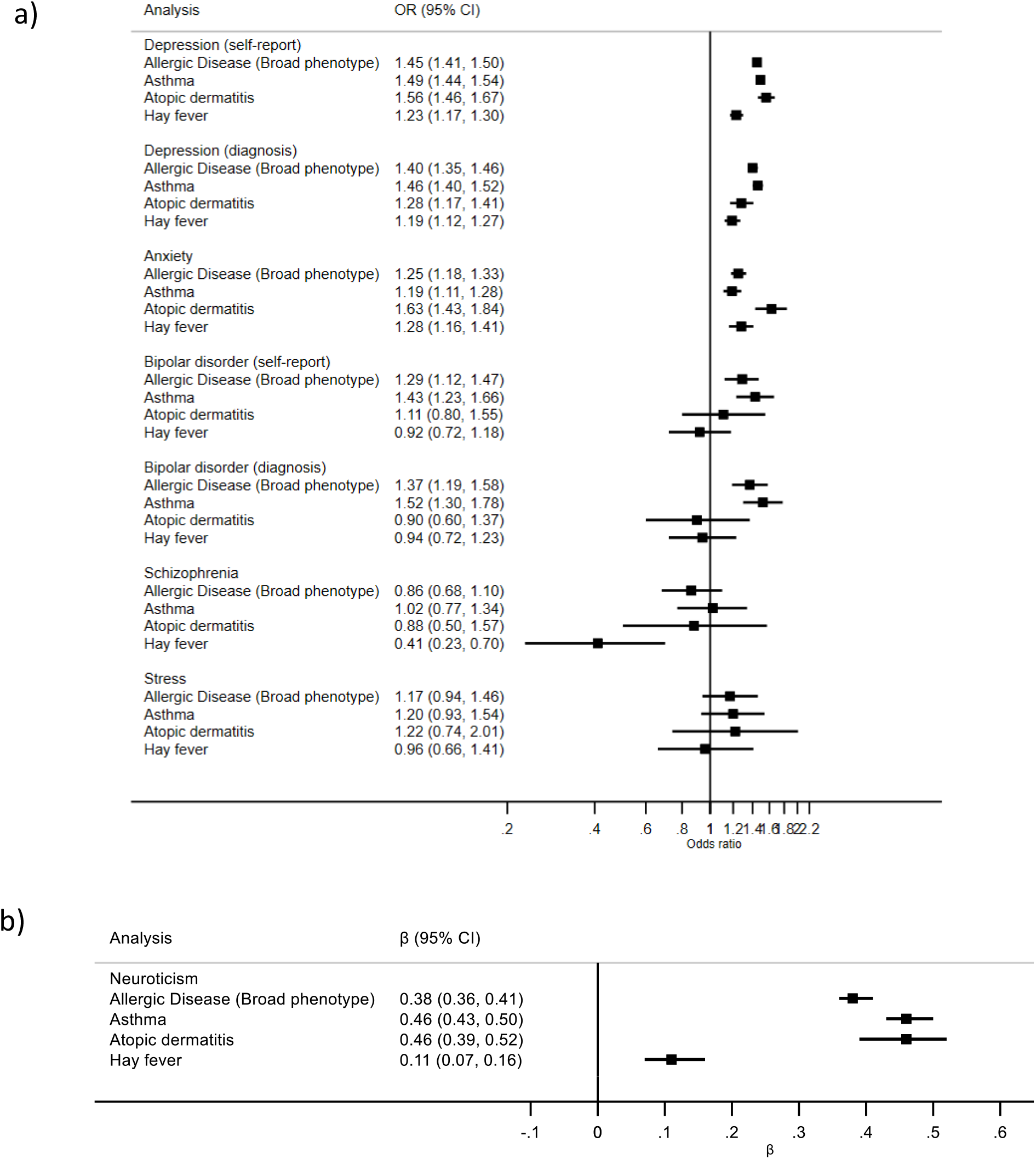
Phenotypic association between broad allergic disease and mental health and personality traits. a) Estimates (ORs) given for odds of mental health traits in allergic disease sufferers versus non-sufferers. b) Neuroticism estimate (β) given for change in summary score in allergic disease sufferers versus non-sufferers. CI, confidence interval.

Anxiety also showed evidence for association with the broad allergic disease phenotype (OR = 1.25; 95% CI = 1.18, 1.33; *P*-value=6.45×10^−13^), where stronger evidence was seen with AD and weaker associations with asthma and hay fever (Figure 2a, see Table E11 in the Online Repository). Bipolar disorder was associated with the broad allergic disease phenotype and appeared to be driven by asthma. This association was not consistent for hay fever or AD. (Figure 2a, see Table E11 in the Online Repository). Of the allergic disease phenotypes investigated only hay fever showed evidence of association with schizophrenia with a protective direction of effect (OR = 0.41; 95% CI = 0.23, 0.70; *P*-value = 1.36×10^−3^) (Figure 2a, see Table E11 in the Online Repository). There was very weak evidence of a phenotypic association between stress and the allergic disease phenotypes. All phenotypes showed evidence of association with neuroticism, most strongly with the broad allergic disease phenotype, asthma and AD (Figure 2b, see Table E11 in the Online Repository).

### MR analyses

#### Causal effect of allergic disease genetic risk upon psychiatric traits – broad phenotype

Two-sample MR found little evidence that genetic liability for the broad allergic disease phenotype causally increases the risk of psychiatric traits (Figure 3a, see Table E12 in the Online Repository). When investigating the causal effect upon MDD risk, the MR estimate was in a consistent direction with that found phenotypically (Figure 2a); however, the phenotypic associations failed to replicate in the MR analysis (OR = 1.01 per doubling odds of allergic disease; 95 % CI = 0.97, 1.06; *P-*value = 0.51). These estimates suggest that a reduction in MDD risk > 21% or increase in risk MDD > 6%, which would be clinically important effects, are unlikely. There was weak evidence that genetic liability for the broad allergic disease phenotype increased the risk bipolar disorder. The direction of effect was consistent with the phenotypic estimate, though the confidence intervals crossed the null line (OR = 1.05; 95% CI = 0.99, 1.12; *P-*value = 0.10) (Figure 3a, see Table E12 in the Online Repository), suggests a reduction in risk > 1% or increase > 12% is unlikely. The magnitude of effect was also smaller than that found phenotypically.

**Figure 3.**
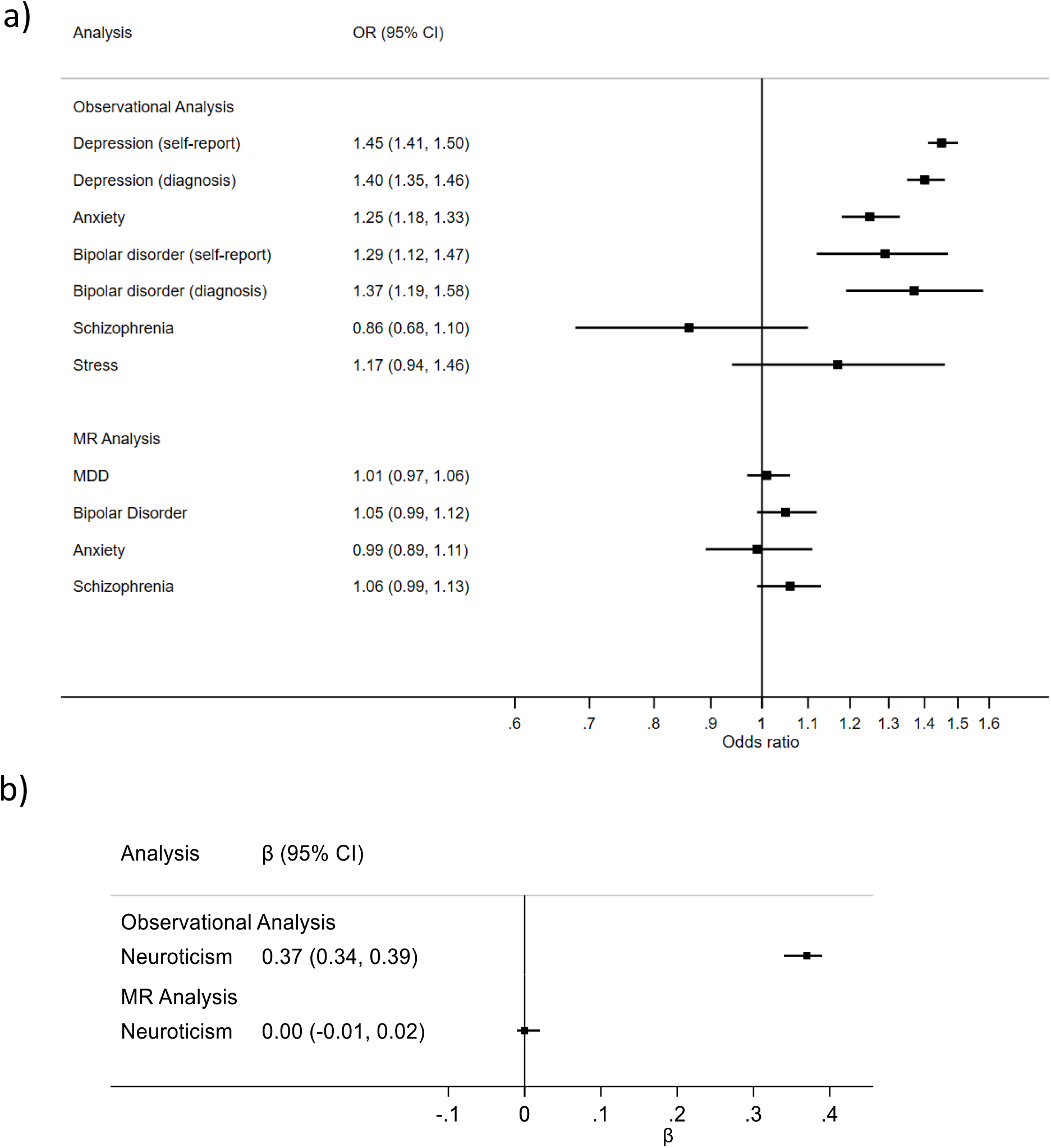
Causal effect of broad allergic disease genetic liability upon mental health outcomes. a) Observational analysis: Estimates (ORs) are given for odds of mental health outcomes in individuals with the broad allergic disease phenotype versus those without. MR Analysis: Estimates are given for odds of mental health outcomes per doubling odds of broad allergic disease phenotype. b) Observational analysis: Neuroticism estimate (β) given for change in summary score in allergic disease sufferers versus non-sufferers. MR Analysis: Causal estimate given for change in total neuroticism score per doubling odds of broad allergic disease phenotype. CI, confidence interval

When looking at the causal effects on anxiety, schizophrenia and neuroticism, the phenotypic association we observed with the broad allergic disease phenotype failed to replicate, and causal effect estimates were null (Figure 3).

#### Causal effect of allergic disease genetic risk upon psychiatric traits – individual phenotypes

When assessing the individual allergic disease phenotypes, the strongest effect was found between hay fever and increased risk of bipolar disorder (OR = 1.09; 95% CI = 1.00, 1.19; *P-*value = 0.06) (see Figure E2 in the Online Repository) and effect estimates were consistent across each sensitivity analysis (Figure E3 in the Online Repository). Furthermore, no variants were excluded from the hay fever genetic instrument upon Steiger filtering. However, the confidence intervals did cross the null line and there was some heterogeneity detected (Q = 65; *P-*value = 3.69×10^−04^). There was little evidence of a phenotypic association between hay fever and bipolar disorder, and the effects were in opposing directions for self-report and diagnosis.

Although the strongest phenotypic association was identified between bipolar disorder and asthma, we found little evidence that genetic liability for asthma influenced the risk of bipolar disorder (see Figure E4 in the Online Repository) and the confidence intervals did not overlap with the phenotypic estimates (OR = 1.04 per doubling odds of asthma; 95% CI = 0.98, 1.10; *P-*value = 0.19).

While anxiety and neuroticism were phenotypically associated with asthma, AD and hay fever, there was little evidence that these relationships were causal in the MR analysis. The causal effect estimates were close to the null and were generally not in a consistent direction to the phenotypic estimates (see Figure E2, E4, E5 and E6 in the Online Repository).

#### Casual effect of psychiatric disorder genetic risk upon allergic disease

We found little evidence that any of the mental health phenotypes causally affected the broad allergic disease phenotype (see Table E13 in the Online Repository). There was some weak evidence of a causal effect of MDD genetic risk on the broad allergic disease phenotype (OR=1.10 per doubling odds of MDD; 95% CI = 0.98, 1.23; *P-*value = 0.10), however, this was of a much smaller magnitude than the phenotypic association and the confidence intervals did not overlap (see Figure E7 in the Online Repository). There was also weak evidence found for neuroticism genetic risk causally increasing risk for the broad allergic disease phenotype (OR = 1.06 per unit change in total neuroticism score; 95% CI = 1.00, 1.13; *P-*value = 0.05). This again was of a smaller magnitude and had an opposite direction of effect to the phenotypic association (See Table E13 and Figure E8 in the Online Repository). Furthermore, the effect estimates obtained from the sensitivity analyses were not consistent in direction and there was some heterogeneity detected (Q = 65; *P-*value = 1.32×10^−05^) (See Figure E9 in the Online Repository).

For the individual allergic disease phenotypes, we found some evidence for bipolar disorder genetic liability having a protective effect upon hay fever (OR=0.94 per doubling odds of bipolar disorder; 95 % CI = 0.90, 0.99; *P-*value = 0.02) (see Figure E10 in the Online Repository); which is weaker than the magnitude of effect found for hay fever genetic risk upon bipolar disorder. Estimates obtained from the MR sensitivity analyses also had a protective direction of effect (see Figure E11 in the Online Repository), with little evidence of horizontal pleiotropy (MR-Egger intercept = 0.03; 95% CI = −0.02, 0.08; *P-*value = 0.21) or heterogeneity (Q = 7.4; *P* = 0.99). Steiger filtering did not exclude any SNPs from the bipolar disorder instrument. However, a protective effect of bipolar disorder upon hay fever was not observed in the phenotypic analysis.

The phenotypic associations between depression and anxiety with asthma, AD and hay fever individually were not replicated in the MR analyses of mental health traits upon these allergic disease phenotypes. This was also seen for the association of bipolar disorder with AD and asthma, and also between schizophrenia and hay fever (see Figure E10, E12, E13 in the Online Repository).

## Discussion

We investigated the association between allergic disease and mental health using both observational multivariable adjusted regression based on UK Biobank data, and two-sample MR approaches based on publicly available GWAS summary data. With the exception of schizophrenia and stress, we found strong evidence of phenotypic associations between all mental health and personality phenotypes investigated with the broad allergic disease phenotype, particularly with depression, replicating findings previously reported in a Taiwanese population^1^. We also identified associations with specific allergic diseases (hay fever, AD, asthma). Anxiety and neuroticism were phenotypically associated with each of the individual diseases, but the association between the broad allergic disease phenotype and bipolar disorder appeared to be driven by asthma. However, when using an MR approach, we found very little evidence that any of these phenotypic associations were likely to be causal, and where there was evidence for this, the causal effect estimates were of a much smaller magnitude. Given these results, it seems likely that the majority of phenotypic associations between allergic disease and mental health are due to confounding or some other form of bias. This suggests that the observed clinical co-morbidity between allergic disease and mental health problems is unlikely to be causal and that intervening to prevent onset of allergic disease is unlikely to directly improve mental health (and vice versa).

We did find some evidence of a causal effect of genetic liability for bipolar disorder upon hay fever risk. However, we did not find evidence for a phenotypic association. It is interesting to note that Steiger filtering gave evidence that the correct direction of causality was being tested in both instances. Although evidence for a casual effect of hay fever genetic risk upon bipolar disorder was weak, it is possible that this relationship acts through inflammatory mechanisms. This would support the inflammatory hypothesis, where several psychiatric traits, including bipolar disorder have been reported to be associated with increased inflammatory markers^11^.

Although our findings suggest that there is no direct causal effect of onset of allergic disease on the onset of mental health phenotypes, there is some evidence that effectively treating skin disease can improve mental wellbeing^39^. A recent report by the All-Party Parliamentary Group on Skin (APPGS) has recommended that patients with a chronic skin condition should receive an annual assessment of the psychological impact of their condition^40^. In our study the genetic instruments we used were identified in GWAS which looked at the onset of allergic disease or mental health phenotypes. In primary care and trial settings, interventions often aim to improve these phenotypes rather than prevent their onset, which could have different causal relationships^13^. Methods are currently being developed which will enable us to investigate the causal effects of disease progression rather than disease onset, such as the recent slope hunter approach proposed by Mahmood and colleagues^41^. Future work should follow up these relationships to investigate whether interventions that aim to improve allergic disease show stronger evidence of a causal effect on mental health (and vice versa).

Triangulating findings using both observational and MR approaches using studies with large sample sizes, and across multiple contexts by comparison with published findings from international cohorts, is a particular strength of this study. This enables us to compare the magnitude of effect across approaches. For the majority of our analyses, the confidence intervals for the observational and causal effect estimates did not overlap, indicating that any causal effect of allergic disease is likely to be much smaller than the phenotypic estimates suggest. Although we did not find strong evidence for a clinically relevant causal effect of allergic disease on mental health (or vice versa), the MR estimates particularly for the allergic disease exposures are precise, enabling us to estimate the maximum causal effect we are likely to see. In addition, investigating both the broad allergic disease phenotype and separating out the specific effects of asthma/AD/hay fever, enabled us to determine whether the relationship is driven by certain phenotypes.

There are some limitations to this study that should be considered. First, the available instruments for the mental health phenotypes were weaker than those used for the allergic disease phenotypes. It is therefore difficult to rule out bidirectional effects that were not uncovered, as it is possible that we did not have enough power to detect causal effects of mental health and personality on allergic disease. Second, although the evidence was strongest for the causal effect of bipolar disorder genetic liability upon hay fever, after accounting for multiple testing using a Bonferroni correction, none of the MR analyses passed the suggested threshold (*P* < 1.25×10^−3^). However, this approach is likely overly conservative given the correlation between our phenotypes. Third, the UK Biobank phenotypes used within this study were predominantly self-reported, which could increase the potential for misreporting. However, where possible we repeated the observational analysis using mental health diagnoses, and the results were consistent. Fourth, there was some sample overlap in our MR analyses of neuroticism and both the broad allergic disease and hay fever phenotype, due to the inclusion of UK Biobank in both GWAS. Sample overlap can bias the causal estimate towards the phenotypic exposure-outcome association^31^, however our MR estimates for these associations were null so this was not an issue. Fifth, the results from this study may not be generalisable across populations. Both the observational data and the publicly available GWAS data used for the MR used data from predominantly white European samples in adulthood. More diverse GWAS are required to improve transferability of results across populations and age groups^42^. Finally, it is important to note that here we only estimate the effects of genetic liability to a particular phenotype, not the effect of developing the phenotype^29^. It is therefore possible that getting specific allergic disease or mental health phenotypes could have different effects to those reported here.

In conclusion, few of the observed associations between allergic disease and mental health were replicated. The causal effect we did identify appears to be much lower in magnitude than that suggested observationally. This suggests that the majority of co-incidence observed clinically is unlikely to be causal. Therefore, intervening to prevent onset of allergic disease in unlikely to directly prevent the onset of mental ill-health. But future work should aim to investigate whether interventions that aim to improve allergic disease have a causal effect on mental health (and vice versa).

## Supporting information

Supplementary tables - genetic instruments

## Data Availability

Data from UK Biobank (as used in observational analyses) are available to researchers on request, subject to a data access fee. The code and datasets used to carry out the MR analyses are available on GitHub (https://github.com/abudu-aggrey/Allergic_Disease_Mental_Health_MR).

## Acknowledgements

This research has been conducted using the UK Biobank Resource under Application Number 9142. Details of patient and public involvement in the UK Biobank are available online (http://www.ukbiobank.ac.uk/about-biobank-uk/ and https://www.ukbiobank.ac.uk/wp-content/uploads/2011/07/Summary-EGF-consultation.pdf). No patients were specifically involved in setting the research question or the outcome measures, nor were they involved in developing plans for recruitment, design, or implementation of this study. No patients were asked to advise on interpretation or writing up of results. There are no specific plans to disseminate the results of the research to study participants, but the UK Biobank disseminates key findings from projects on its website.

## Contributors

AB-A, SJB, LP and HS conceived the study concept. AB-A and HS managed the project. SJ, AB-A and HS performed the statistical analysis. SJ, AB-A, SJB, LP and HS drafted the manuscript. All authors were involved in the interpretation of the data and contributed to and approved the final version of the manuscript.

## Data sharing

The UK Biobank dataset used to conduct the research in this paper is available via application directly to the UK Biobank. Applications are assessed for meeting the required criteria for access, including legal and ethics standards. More information regarding data access can be found here: http://www.ukbiobank.ac.uk/scientists-3/. The code and datasets used to carry out the MR analyses are available on GitHub (https://github.com/abudu-aggrey/Allergic_Disease_Mental_Health_MR).

## Abbreviations

AD: Atopic Dermatitis
CI: Confidence interval
GWAS: Genome-wide association study
IVW: Inverse-variance weighted
MBE: Mode-based estimate
MDD: Major Depressive Disorder
MR: Mendelian Randomization
OR: Odds ratio
SNP: Single-nucleotide polymorphism

