## Supplementary information for "Investigating the causal relationship between allergic disease and mental health"

**Budu-Aggrey, A et al**

**Supplementary Information**

**Table E10 - Summary of genetic instruments and GWAS data used to perform MR analyses**

| **Phenotype** | **Number of variants in instrument** | **N [instrument]**  **(cases/controls)** | **N**  **[summary data]** | **N**  **(cases)** | **N**  **(controls)** | **Reference** |
| --- | --- | --- | --- | --- | --- | --- |
| Allergic Disease | 89 | 360,838  (180,129 /180,709) | 242,569 ^a^ | 96,794 | 145,775 | Ferreira et al, 2017 |
| Asthma | 16 | 127,669 | 127,669 | 19,954 | 107,715 | Demenais et al, 2018 |
| Atopic Dermatitis | 23 | 103,066  (18,900 / 84,166) | 40,835^a^ | 10,788 | 30,047 | Paternoster et al, 2015 |
| Hay Fever | 37 | 212,120  (59,762 / 152,358) | 38,838 ^a,b^ | 10,582 | 28,256 | Waage et al, 2018 |
| Depression (MDD) | 32 | 480,359  (135,458 / 344,901) | 480,359 | 135,458 | 344,901 | Wray et al, 2018 |
| Depression (MDD)^a,b^ | 31 | 143,265 ^a,b^  (45,591 / 97,674) | 143,265 ^a,b^ | 45,591 | 97,674 | Wray et al, 2018 |
| Bipolar Disorder | 22 | 198,882  (29,764 / 169,118) | 51,710 | 20,352 | 31,358 | Stahl et al, 2018 |
| Anxiety | 5 | 83,548  (25,435 / 58,113) | - | - | - | Purves et al, 2019 |
| Anxiety^c^ | - | - | 17,310^b^ | 5,300* | 12,000* | Otowa, 2016 |
| Schizophrenia | 79 | 150,064  (36,989 / 113,075) | 150,064 | 36,989 | 113,075 | Ripke et al, 2014 |
| Neuroticism | 66 | 329,821 | - | - | - | Luciano M et al, 2017 |

MDD, Multiple depressive disorder; ^a^ Excludes 23andMe individuals; ^b^ Excludes UK Biobank individuals; ^c^ Where full summary statistics were not available for the most recent, largest GWAS, an alternative study (with full summary statistics) was used when the listed phenotype was the outcome in the MR analysis *numbers are approximate

**Table E11 - Observational relationship between allergic disease phenotypes and mental health outcomes**

|  | **Allergic Disease**  **(broad phenotype)** | | | | **Atopic Dermatitis** | | | | **Asthma** | | | | **Hay fever** | | | |
| --- | --- | --- | --- | --- | --- | --- | --- | --- | --- | --- | --- | --- | --- | --- | --- | --- |
|  | **N** | **OR** | **95% CI** | ***P*-value** | **N** | **OR** | **95% CI** | ***P*-value** | **N** | **OR** | **95% CI** | ***P*-value** | **N** | **OR** | **95% CI** | ***P*-value** |
| Depression (self-report) | 448,634 | 1.45 | 1.41, 1.50 | 3.6x10^-130^ | 452,609 | 1.56 | 1.46, 1.67 | 7.1x10^-40^ | 451,332 | 1.49 | 1.44, 1.54 | 1.7x10^-112^ | 452,691 | 1.23 | 1.17, 1.30 | 1.8x10^-16^ |
| Depression (diagnosis) | 105,616 | 1.40 | 1.35, 1.46 | 2.6x10^-76^ | 106,471 | 1.28 | 1.17, 1.41 | 2.0x10^-07^ | 106,260 | 1.46 | 1.40, 1.52 | 4.5x10^-74^ | 106,477 | 1.19 | 1.12, 1.27 | 6.6x10^-08^ |
| Anxiety | 448,634 | 1.25 | 1.18, 1.33 | 6.5x10^-13^ | 452,609 | 1.63 | 1.43, 1.84 | 3.3x10^-14^ | 451,332 | 1.19 | 1.11, 1.28 | 2.1x10^-06^ | 452,691 | 1.28 | 1.16, 1.41 | 5.6x10^-07^ |
| Bipolar Disorder  (self-report) | 448,634 | 1.29 | 1.12, 1.47 | 2.8x10^-04^ | 452,609 | 1.11 | 0.80, 1.55 | 0.543 | 451,332 | 1.43 | 1.23, 1.66 | 4.6x10^-06^ | 452,691 | 0.92 | 0.72, 1.18 | 0.533 |
| Bipolar disorder (diagnosis) | 78,617 | 1.37 | 1.19, 1.58 | 1.1x10^-05^ | 79,194 | 0.90 | 0.60, 1.37 | 0.630 | 79,084 | 1.52 | 1.30, 1.78 | 1.1x10^-07^ | 79,201 | 0.94 | 0.72, 1.23 | 0.655 |
| Schizophrenia | 448,634 | 0.86 | 0.68, 1.10 | 0.238 | 452,609 | 0.88 | 0.50, 1.57 | 0.675 | 451,332 | 1.02 | 0.77, 1.34 | 0.904 | 452,691 | 0.41 | 0.23, 0.70 | 0.001 |
| Stress | 448,634 | 1.17 | 0.94, 1.46 | 0.150 | 452,609 | 1.22 | 0.74, 2.01 | 0.435 | 451,332 | 1.20 | 0.93, 1.54 | 0.159 | 452,691 | 0.96 | 0.66, 1.41 | 0.838 |
|  | **N** | **Beta** | **95% CI** | ***P*-value** | **N** | **Beta** | **95% CI** | ***P*-value** | **N** | **Beta** | **95% CI** | ***P*-value** | **N** | **Beta** | **95% CI** | ***P*-value** |
| Neuroticism | 364,265 | 0.38 | 0.36, 0.41 | 6.8x10^-166^ | 367,334 | 0.46 | 0.39, 0.52 | 5.9x10^-42^ | 366,501 | 0.46 | 0.43, 0.50 | 2.4x10^-174^ | 367,398 | 0.11 | 0.07, 0.16 | 8.1x10^-07^ |

Univariable analysis - adjusted for age and sex. CI, confidence interval; OR, odds ratio

**Table E12 - MR analyses for causal effect of allergic disease genetic liability on mental health outcomes**

| **Analysis** | **Causal Estimate (95% CI)** | ***P*-value** |
| --- | --- | --- |
| Allergic Disease – MDD | 1.01 (0.97, 1.06) | 0.51 |
| Allergic Disease – Bipolar Disorder | 1.05 (0.99, 1.12) | 0.10 |
| Allergic Disease – Anxiety | 0.99 (0.89, 1.11) | 0.90 |
| Allergic Disease - Schizophrenia | 1.06 (0.99, 1.13) | 0.08 |
| Allergic Disease – Neuroticism* | 0.01 (-0.01, 0.03) | 0.38 |
| Asthma – MDD | 0.99 (0.96, 1.03) | 0.71 |
| Asthma – Bipolar Disorder | 1.04 (0.98, 1.10) | 0.19 |
| Asthma – Anxiety | 0.98 (0.88, 1.10) | 0.76 |
| Asthma - Schizophrenia | 1.01 (0.95, 1.07) | 0.78 |
| Asthma – Neuroticism* | 0.00 (-0.02, 0.03) | 0.90 |
| Atopic Dermatitis – MDD | 1.02 (0.97, 1.07) | 0.50 |
| Atopic Dermatitis – Bipolar Disorder | 1.04 (0.96, 1.13) | 0.34 |
| Atopic Dermatitis – Anxiety | 1.06 (0.97, 1.16) | 0.19 |
| Atopic Dermatitis - Schizophrenia | 1.01 (0.93, 1.10) | 0.73 |
| Atopic Dermatitis – Neuroticism* | 0.01 (-0.02, 0.03) | 0.64 |
| Hay fever – MDD | 1.00 (0.94, 1.07) | 0.98 |
| Hay fever – Bipolar Disorder | 1.09 (1.00, 1.19) | 0.06 |
| Hay fever – Anxiety | 1.00 (0.90, 1.12) | 0.95 |
| Hay fever - Schizophrenia | 1.05 (0.97, 1.14) | 0.23 |
| Hay fever – Neuroticism* | -0.01 (-0.04, 0.01) | 0.22 |

Estimates given for odds of mental health outcome per doubling odds of allergic disease phenotype

*Estimates given for change in total neuroticism score per doubling odds of allergic disease phenotype.

CI, confidence interval; MDD, major depressive disorder

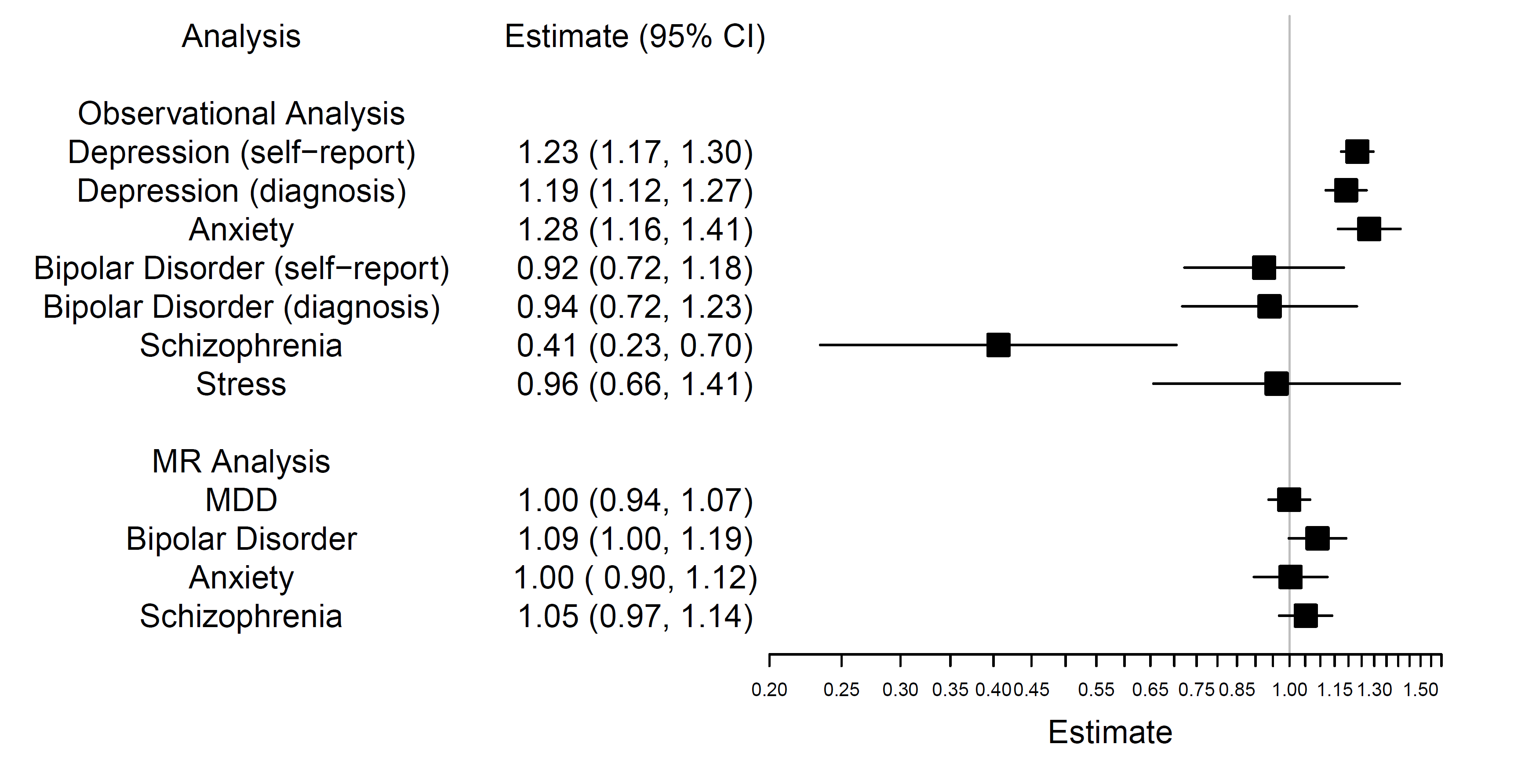

**Figure E2 - Causal effect of hay fever genetic liability upon mental health outcomes**

Observational analysis: Estimates are given for odds of mental health outcomes in individuals with hay fever versus those without. MR Analysis: Estimates are given for odds of mental health outcomes per doubling odds of hay fever.

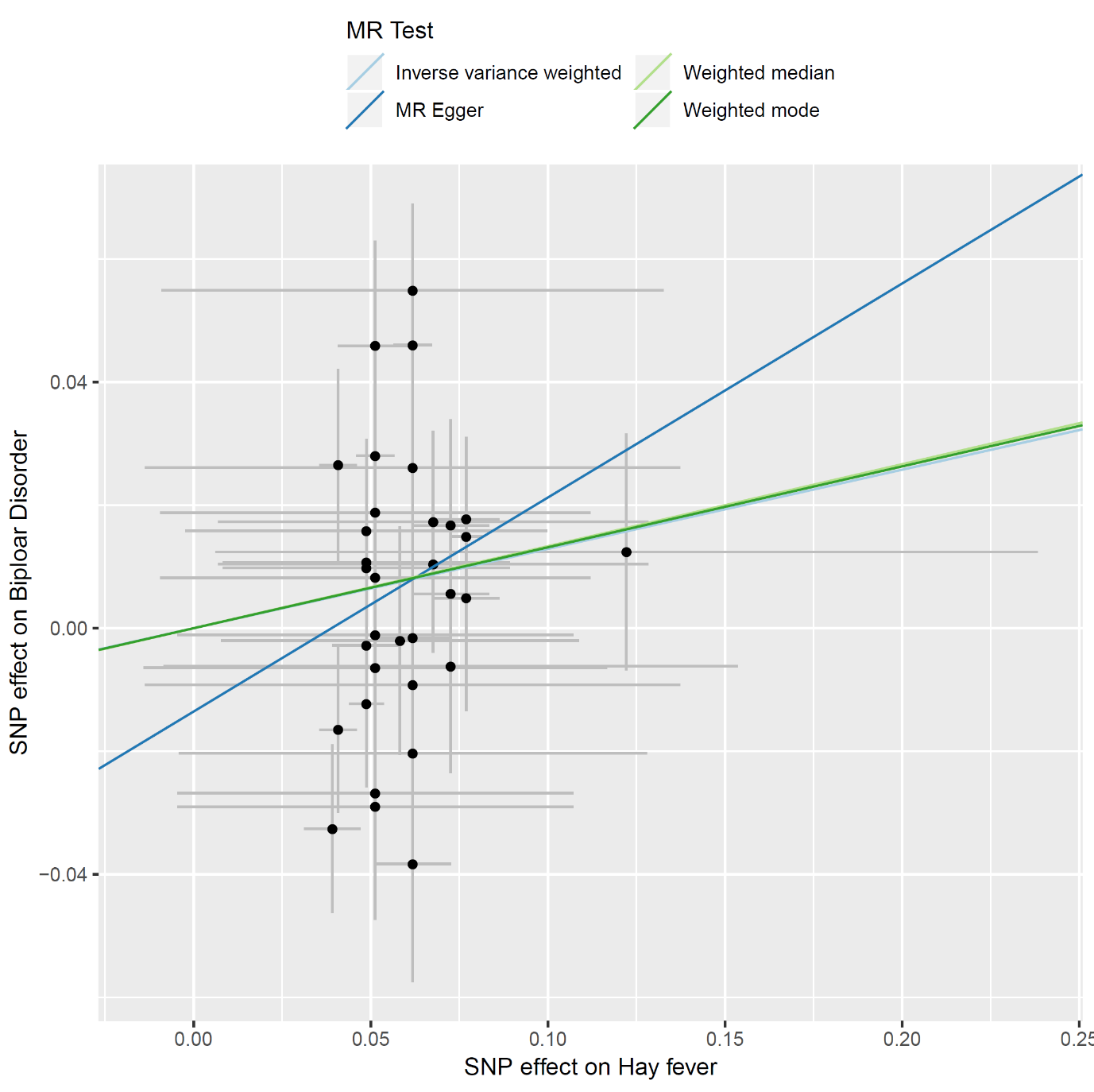

**Figure E3:** **Association of Hay Fever SNPs with Bipolar Disorder**

Estimates are plotted for ln(OR). MR Egger Estimate: 0.34 (95% CI = -0.21 to 0.89); weighted median estimate: 0.11 (95% CI = -0.03 to 0.26); weighted mode estimate: 0.12 (95% CI = -0.09 to 0.32); Egger intercept: -0.01 (95% CI = -0.05 to 0.02); Q statistic: 65 (*P*-value= 3.69×10^-04^).

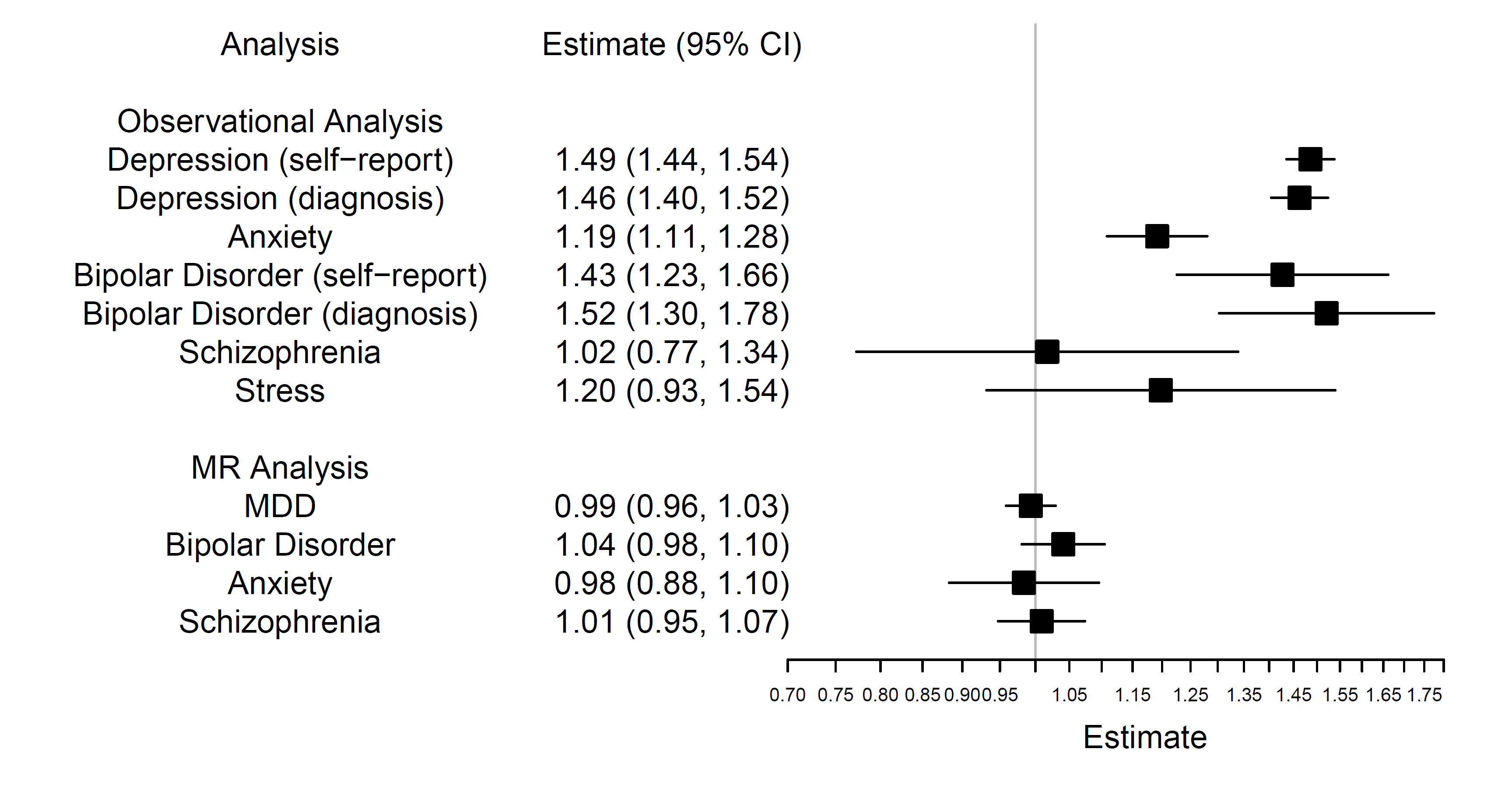

**Figure E4 - Causal effect of asthma genetic liability upon mental health outcomes**

Observational analysis: Estimates are given for odds of mental health outcomes in individuals with asthma versus those without. MR Analysis: Estimates are given for odds of mental health outcomes per doubling odds of asthma.

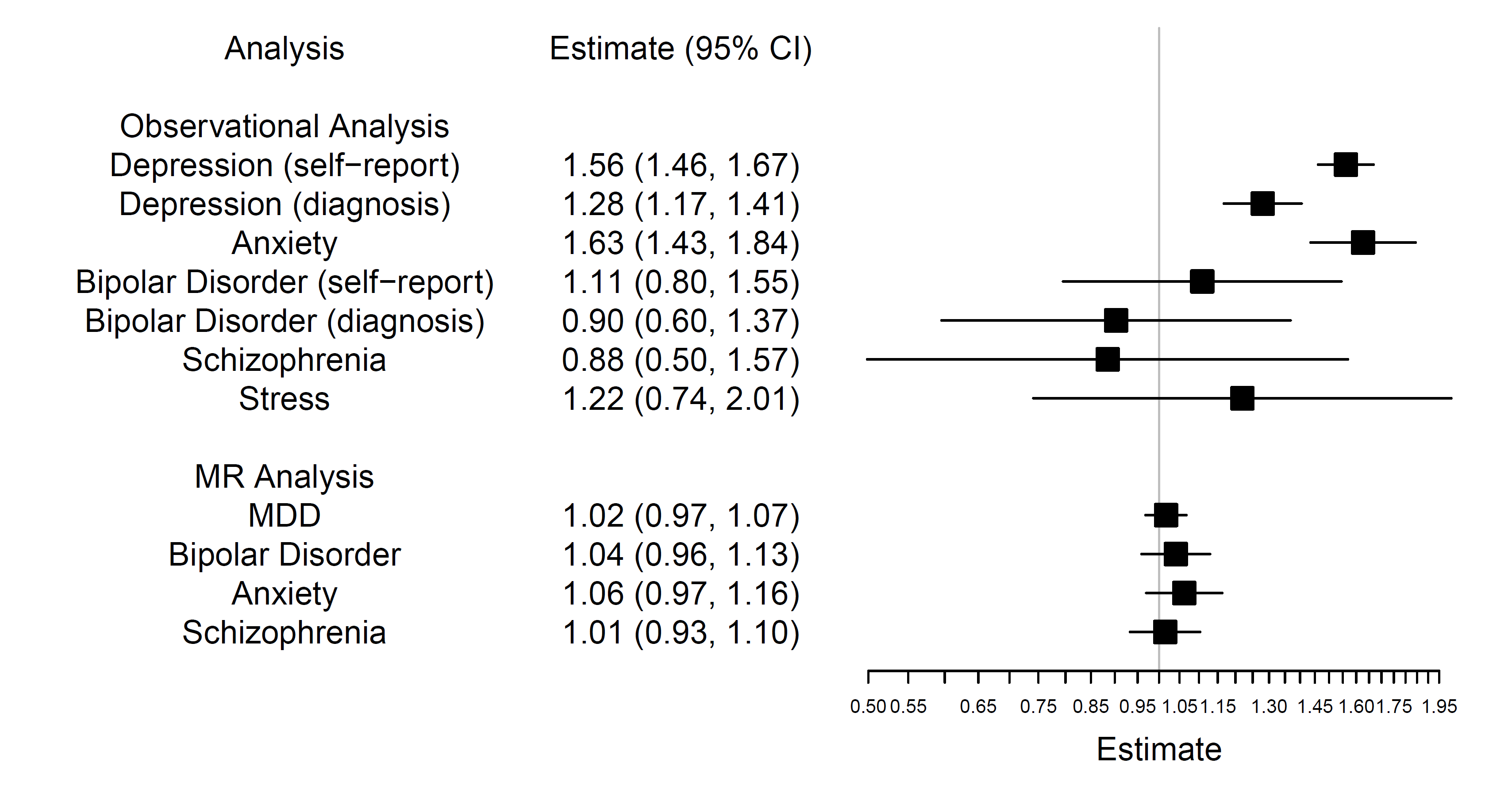

**Figure E5 - Causal effect of atopic dermatitis (AD) genetic liability upon mental health outcomes**

Observational analysis: Estimates are given for odds of mental health outcomes in individuals with AD versus those without. MR Analysis: Estimates are given for odds of mental health outcomes per doubling odds of AD.

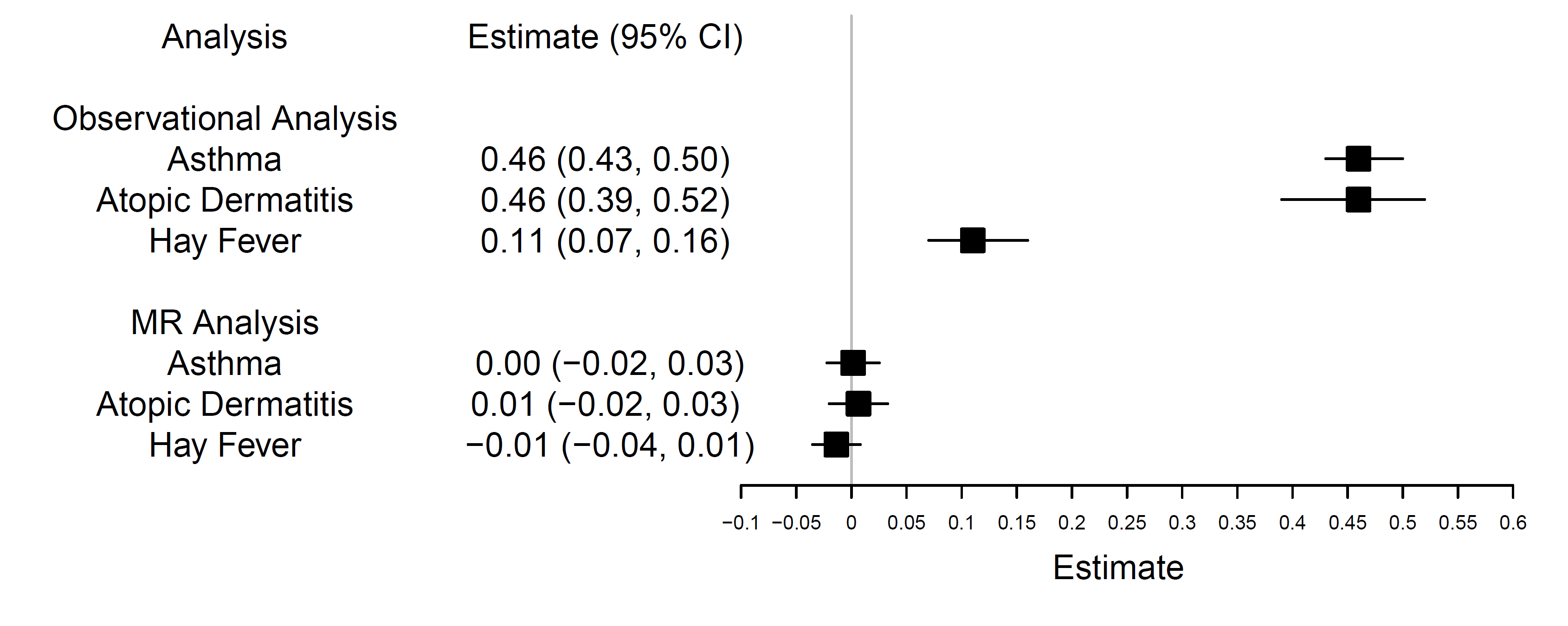

**Figure E6 – Causal effect of asthma, atopic dermatitis (AD) and hay fever liability upon neuroticism**

Observational analysis: Neuroticism estimate given for change in summary score in allergic disease sufferers versus non-sufferers. MR Analysis: Causal estimate given for change in total neuroticism score per doubling odds of allergic disease. CI, confidence interval

**Table E13 - MR analyses for causal effect of genetically predicted mental health outcomes on allergic disease phenotypes**

| **Analysis** | **Causal Estimate (95% CI)** | ***P*-value** |
| --- | --- | --- |
| MDD – Allergic Disease | 1.10 (0.98, 1.23) | 0.10 |
| MDD – Asthma | 1.18 (0.91, 1.53) | 0.22 |
| MDD – Atopic Dermatitis | 1.00 (0.82, 1.21) | 0.96 |
| MDD – Hay Fever | 1.13 (0.97, 1.33) | 0.13 |
| Bipolar Disorder – Allergic Disease | 0.99 (0.95, 1.03) | 0.58 |
| Bipolar Disorder – Asthma | 1.05 (0.93, 1.19) | 0.42 |
| Bipolar Disorder – Atopic Dermatitis | 0.96 (0.89, 1.03) | 0.21 |
| **Bipolar Disorder – Hay Fever** | **0.94 (0.90, 0.99)** | **0.02** |
| Anxiety – Allergic Disease | 1.02 (0.92, 1.12) | 0.77 |
| Anxiety – Asthma | 1.01 (0.95, 1.08) | 0.68 |
| Anxiety – Atopic Dermatitis | 1.04 (0.94, 1.15) | 0.46 |
| Anxiety – Hay Fever | 0.99 (0.91, 1.07) | 0.77 |
| Schizophrenia – Allergic Disease | 1.01 (0.99, 1.03) | 0.39 |
| Schizophrenia – Asthma | 1.01 (0.95, 1.07) | 0.72 |
| Schizophrenia – Atopic Dermatitis | 0.99 (0.95, 1.04) | 0.79 |
| Schizophrenia – Hay Fever | 1.01 (0.96, 1.05) | 0.77 |
| Neuroticism*– Allergic Disease | 1.06 (1.00, 1.13) | 0.05 |
| Neuroticism*– Asthma | 1.12 (0.96, 1.30) | 0.16 |
| Neuroticism*– Atopic Dermatitis | 0.97 (0.82, 1.14) | 0.71 |
| Neuroticism*– Hay Fever | 0.97 (0.85, 1.12) | 0.70 |

Estimates given for odds of allergic disease phenotype per doubling odds of mental health trait

*Estimates given for odds of allergic disease phenotype per unit change in total neuroticism score Estimates with *P*-value < 0.05 are highlighted in **bold**

CI, confidence interval; MDD, major depressive disorder

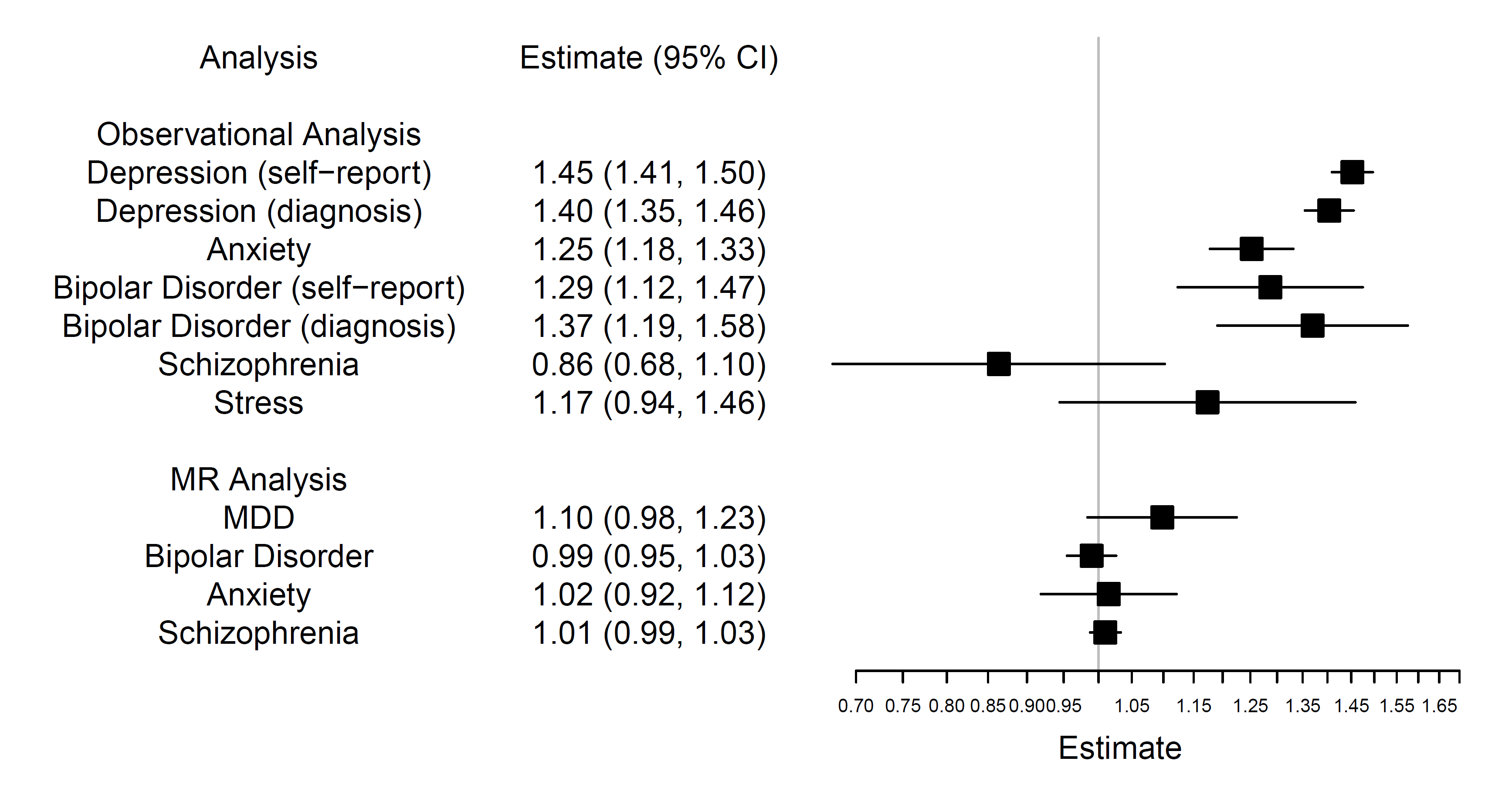

**Figure E7 - Causal effect of mental health genetic liability upon the broad allergic disease phenotype**

Observational analysis: Estimates are given for odds of mental health outcomes in individuals with the broad allergic disease phenotype versus those without. MR Analysis: Estimates are given for odds of the broad allergic disease phenotype per doubling odds of mental health trait. CI; confidence interval

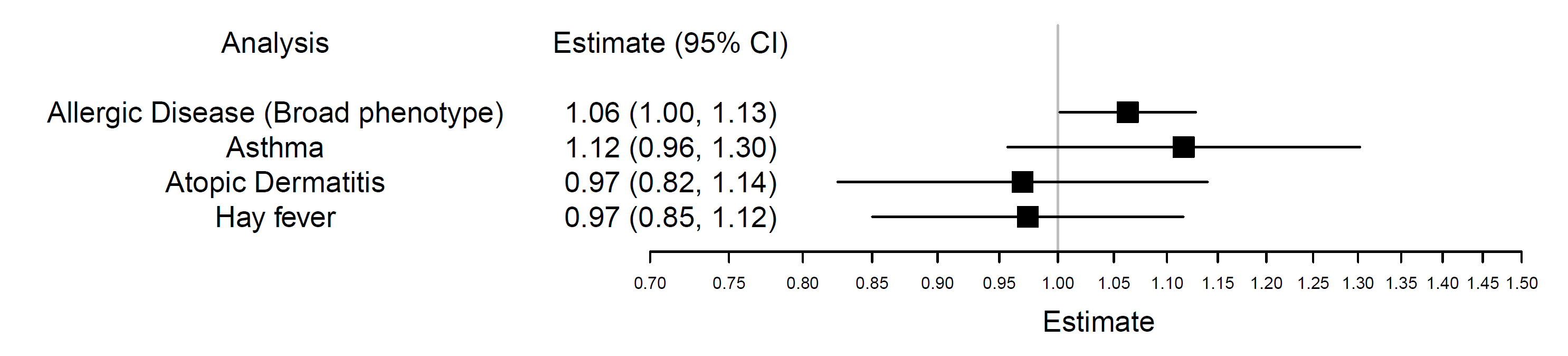

**Figure E8 – Causal effect of neuroticism genetic liability upon allergic disease phenotypes**

Causal estimate given for odds of allergic disease per unit change in total neuroticism score.

CI, confidence interval

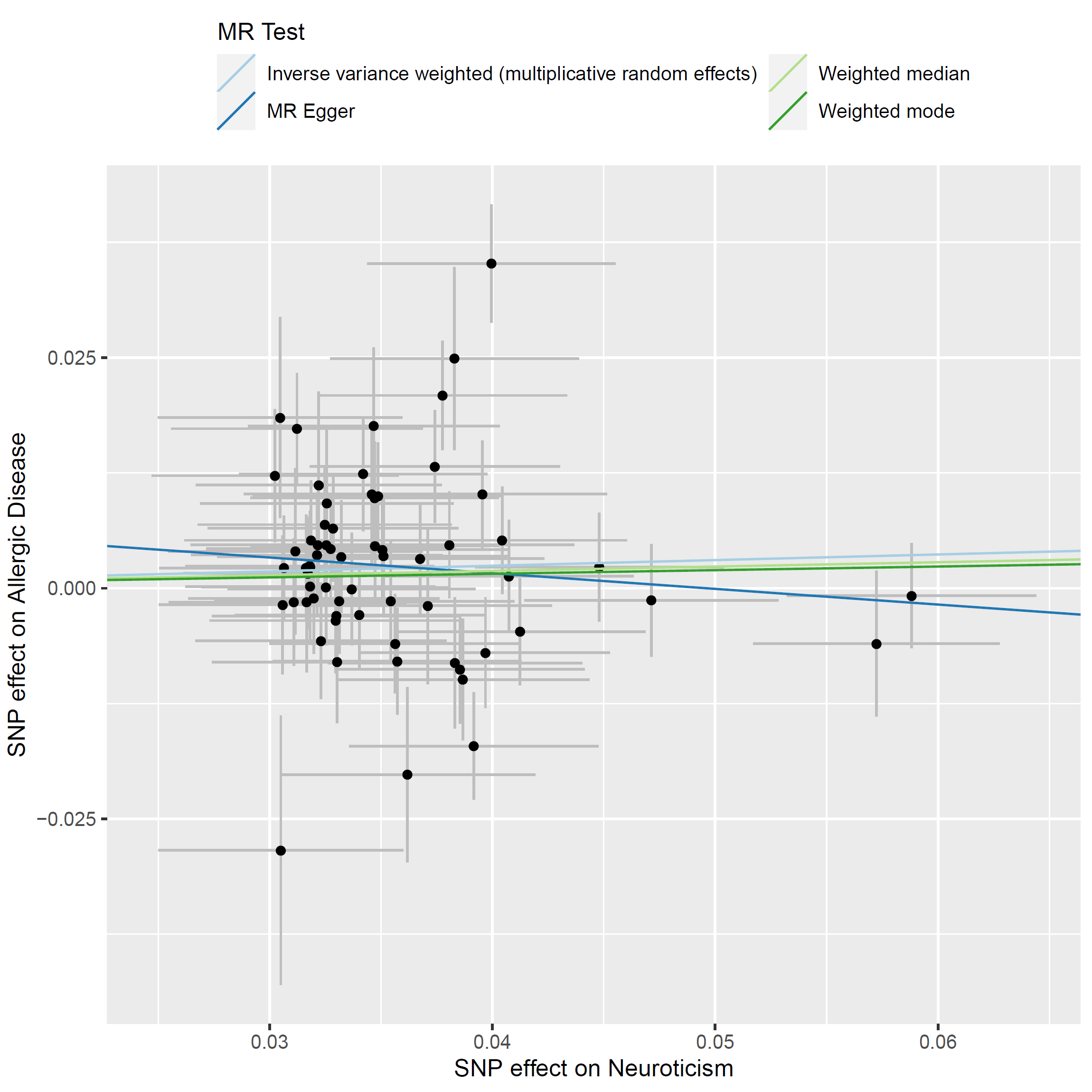

**Figure E9:** **Association of Neuroticism SNPs with the broad allergic disease phenotype**

Estimates are plotted for ln(OR). MR Egger Estimate: -0.17 (95% CI = -0.57 to 0.23); weighted median estimate: 0.05 (95% CI = -0.02 to 0.11); weighted mode estimate: 0.04 (95% CI = -0.08 to 0.16); Egger intercept: 0.01 (95% CI = -0.01 to 0.02); Q statistic: 123 (*P*-value= 1.32×10^-05^).

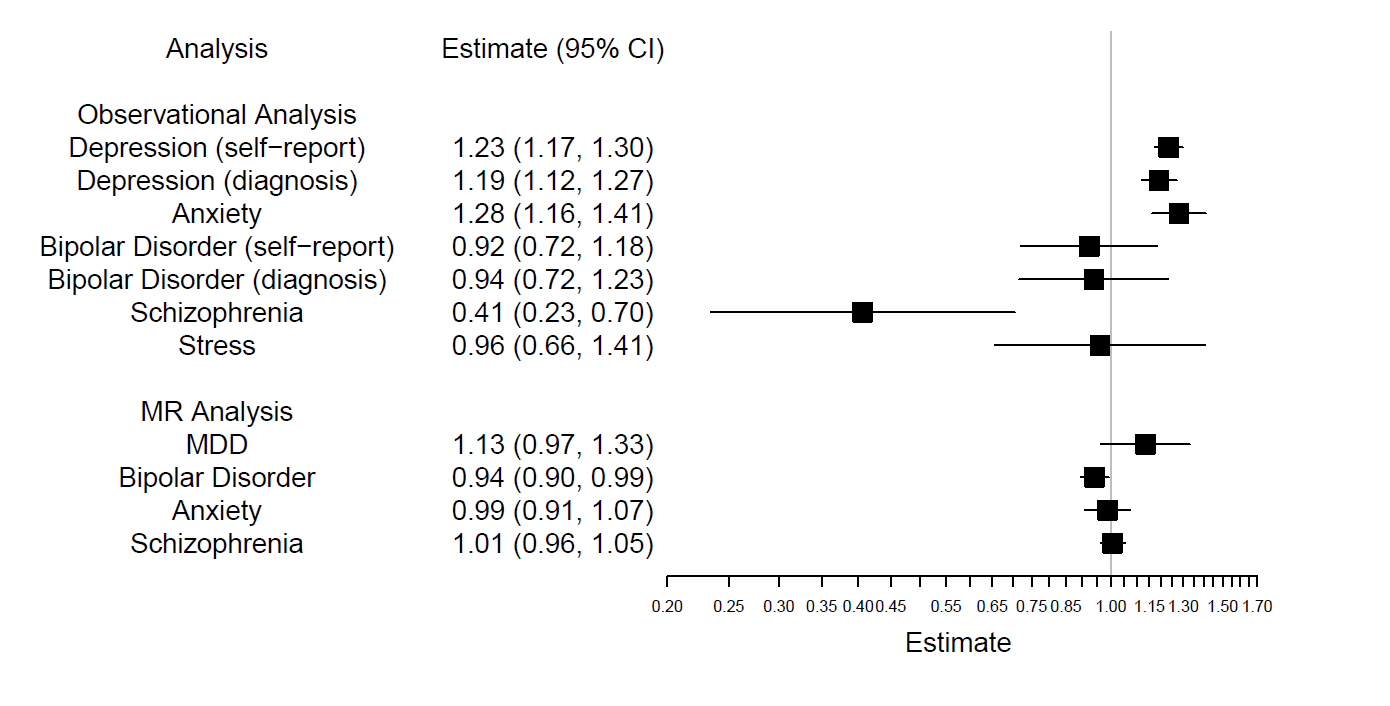

**Figure E10 - Causal effect of mental health genetic liability upon hay fever**

Estimates are given for odds of mental health outcomes in individuals with hay fever versus those without. MR Analysis: Estimates are given for odds of hay fever per doubling odds of mental health trait. CI; confidence interval

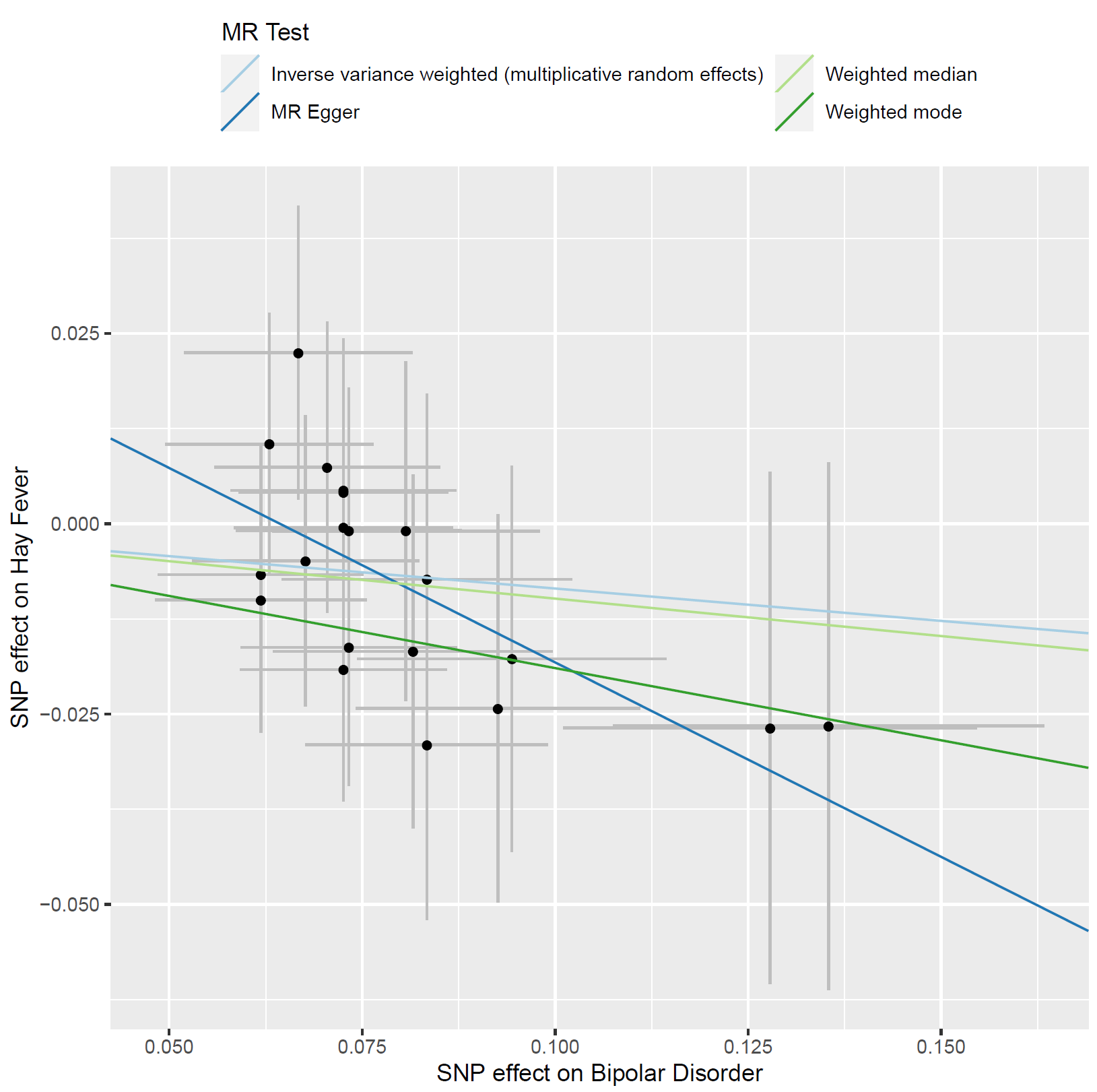

**Figure E11:** **Association of Bipolar Disorder SNPs with Hay Fever**

Estimates are plotted for ln(OR). MR Egger Estimate: -0.51 (95% CI = -1.16 to 0.14); weighted median estimate: -0.10 (95% CI = -0.24 to 0.05); weighted mode estimate: -0.19 (95% CI = -0.47 to 0.10); Egger intercept: 0.03 (95% CI = -0.02 to 0.08); Q statistic: 7.4 (*P*-value= 0.99).

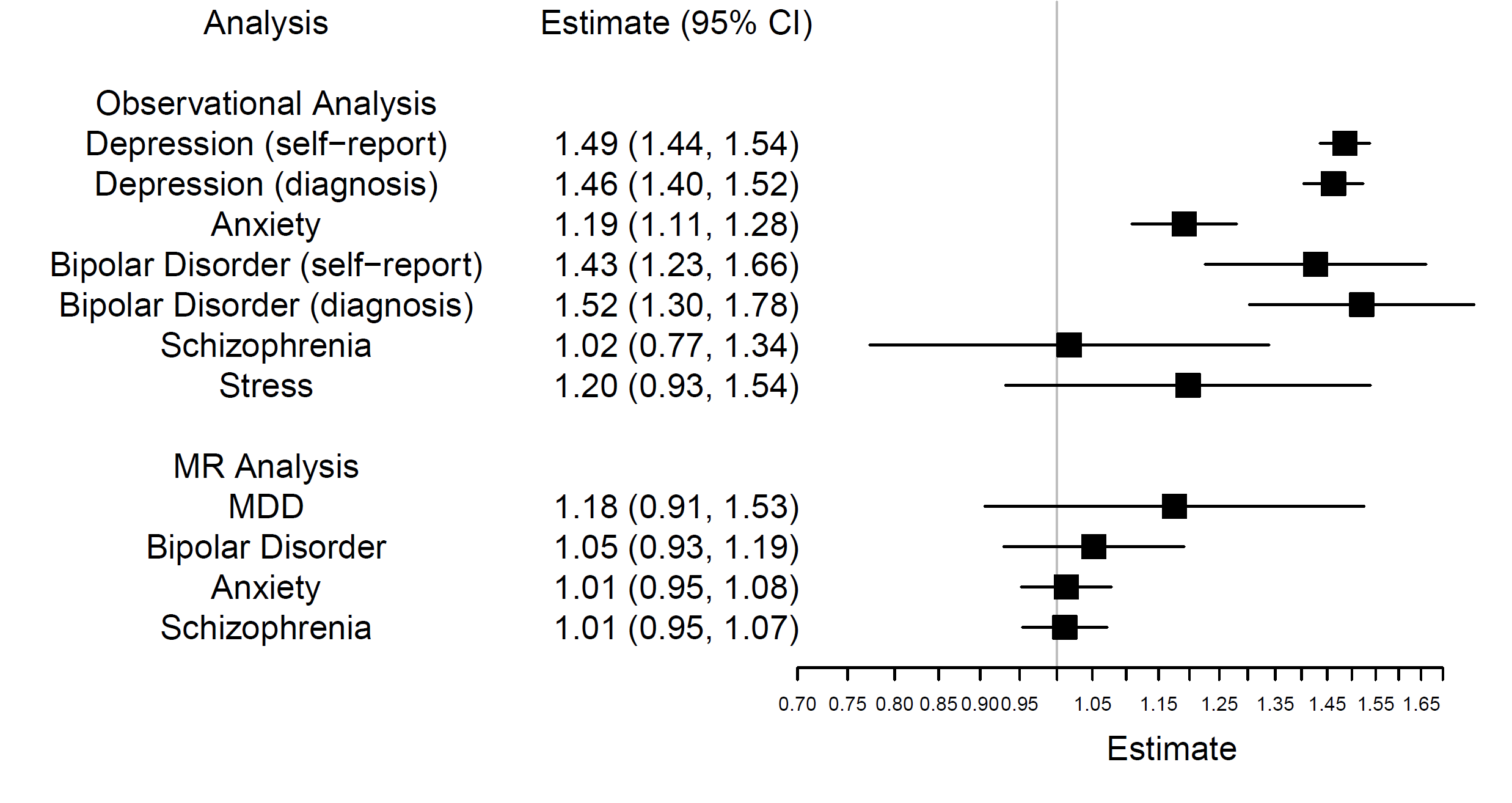

**Figure E12 - Causal effect of mental health genetic liability upon asthma**

Estimates are given for odds of mental health outcomes in individuals with asthma versus those without. MR Analysis: Estimates are given for odds of asthma per doubling odds of mental health trait. CI; confidence interval

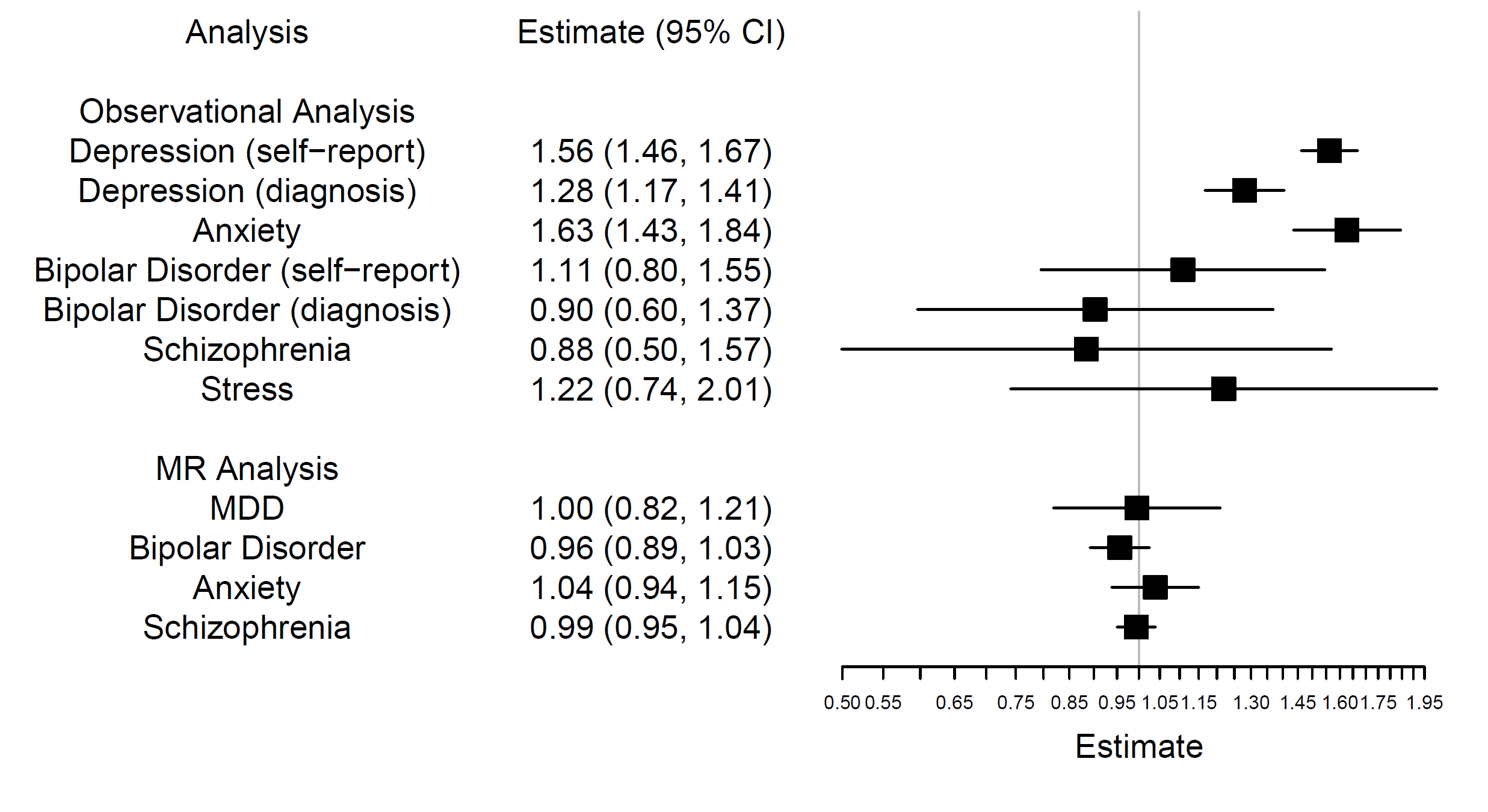

**Figure E13 - Causal effect of mental health genetic liability upon atopic dermatitis (AD)**

Estimates are given for odds of mental health outcomes in individuals with AD versus those without. MR Analysis: Estimates are given for odds of AD per doubling odds of mental health trait. CI; confidence interval
